# Dengue importation into Europe: a network connectivity-based approach

**DOI:** 10.1101/19009589

**Authors:** Donald Salami, César Capinha, Maria do Rosário Oliveira Martins, Carla Alexandra Sousa

**Author notes:** Corresponding author (DS).

## Abstract

The spread of dengue through global human mobility is a major public health concern. A key challenge is understanding the transmission pathways and mediating factors that characterized the patterns of dengue importation into non-endemic areas. Utilizing a network connectivity-based approach, we analyze the importation patterns of dengue fever into European countries.

Seven connectivity indices were developed to characterize the role of the air passenger traffic, seasonality, incidence rate, geographical proximity, epidemic vulnerability, and wealth of a source country, in facilitating the transport and importation of dengue fever. We used generalized linear mixed models (GLMMs) to examine the relationship between dengue importation and the connectivity indices while accounting for the air transport network structure. We also incorporated network autocorrelation within a GLMM framework to investigate the propensity of a European country to receive an imported case, by virtue of its position within the air transport network.

The connectivity indices and dynamical processes of the air transport network were strong predictors of dengue importation in Europe. With more than 70% of the variation in dengue importation patterns explained. We found that transportation potential was higher for source countries with seasonal dengue activity, high passenger traffic, high incidence rates, lower economic status, and geographical proximity to a destination country in Europe. We also found that position of a European country within the air transport network was a strong predictor of the country’s propensity to receive an imported case.

Our findings provide evidence that the importation patterns of dengue into Europe can be largely explained by appropriately characterizing the heterogeneities of the source, and topology of the air transport network. This contributes to the foundational framework for building integrated predictive models for bio-surveillance of dengue importation.

## Introduction

During the last few decades, dengue fever has rapidly spread into new geographical regions with a resultant increase in its global incidence [1, 2]. This global spread has notably been linked to increasing human mobility, particularly air travel [3-5]. Global aviation network has increased in volume by almost eight-fold in the past 40years, enabling human movement across long distances in a relatively short time [6-8]. Thus, creating a mobility network for the spread of infectious diseases like dengue [9-11]. Infected air travelers have contributed significantly to the importation of dengue to non-endemic areas [12-14].

As human mobility and connectivity continue to advance, dengue spread via importation will continue to increase at unpredictable rates [4, 15]. The complexity of the air transport network poses a substantial challenge in the understanding of the dynamics of dengue spread and importation [16, 17]. How to effectively tackle dengue spread mediated via the complex air transport network is a priority of vector-borne disease surveillance and control [18-20]. In this context, understanding the dynamics by which dengue fever is transported, across the complex and dynamic mobility network, is an important first step [5].

Previous work on network-mediated epidemic often assumes a common framework, that the probability of an imported infection is directly correlated with the number of arriving air passengers [13, 21]. However, a simple correlation between imported cases and crude travel statistics is insufficient to explain the transmission pathways [21]. Such an approach does not allow to differentiate imported cases arriving from countries with higher infection risk and transport potential, or if the variance in the number of cases is also mediated by other socio-economic and anthropogenic factors. Neither does this capture the connectivity patterns of the mobility network that influence or constrains the dynamics of importation.

Recent studies have applied a range of social network modeling approaches to understanding the transmission pathways of a network-mediated epidemic. From a general perspective, these methods combine the derived attributes of the epidemic source country and the topology of the transporting network to explain importation dynamics [17, 22]. Epidemic source attributes are characterized by the heterogeneities in the passenger’s air travel volume, socio-economic and anthropogenic factors, that mediate the risk of infection and transport of the disease [23]. While the network topology is characterized by centrality measures of the nodes (countries) in the air transport network. [24, 25]. To our knowledge, there are no studies that apply this explanatory power of social network analysis to characterize dengue importation patterns into Europe.

Here, we adopt a refined network connectivity approach to analyze data on imported dengue cases from 21 European countries, within a 6-year period (2010 – 2015). Specifically, our approach is outlined in the following: (1) We integrate a source-to-destination country combination to construct a network connectivity for dengue importation; (2) We then examine connectivity measures accounting for factors mediating the transport and importation potential from the source country; (3) Lastly, we investigate how the topology of the air transport network influences the importation risk from a source and the propensity of a destination country to receive an imported case.

## Methods

### Conceptual framework

Our proposed network connectivity framework adapts techniques from previous work on dispersal connectivity, spatial autocorrelation and network modelling [25-28] to capture the dynamics of dengue importation. The inputs for our analysis consist of the dengue importation data, air travel data and the underlying air transport network structure.

In the sections below, we describe the various inputs and our modeling approaches. In the first section, we describe the disease and air travel data, with their respective sources. Next, we describe the connectivity indices representing factors that potentially facilitate the transport and importation of dengue from a source country into Europe. We then describe the generalized linear mixed effect (GLMM) modelling approach for quantifying the variation in dengue importation as explained by the indices.

The next section introduces the concept of dependency network analysis to account for the influence of the air transport network structure. Firstly, we describe the construct of the weighted directed network from the air passenger’s data and then define centrality measures to characterize each individual node’s (i.e. country) influence within the network. The centrality measures of the source countries were then added to the GLMM model of the connectivity indices to account for the influence of network structure. Finally, we describe an extension of our analysis to model the propensity of a country in Europe to receive an imported case, by virtue of its network topology (i.e. centrality measures).

### Disease data

We analyzed imported cases of dengue fever reported in Europe for the period of 2010 – 2015. Dengue fever data was obtained from the European Centre for Disease Prevention and Control (ECDC) [29]. Routine (weekly) Europe-wide infectious disease surveillance data is collected from European Economic Area member states (EU/EEA) countries by the ECDC. Data is collected and managed through The European Surveillance System (TESSy) database; a database provided by the ECDC national focal points for surveillance [30].

Here, we considered confirmed cases of dengue, according to the 2012 EU case definition for viral haemorrhagic fever (VHF) which defines a confirmed case as any person meeting the clinical and the laboratory criteria [31]. The subset of imported cases or travel-associated cases are categorized as persons having been outside the country of notification during the incubation period of the disease. Place/country of infection was defined as the place the person was during the incubation period of the disease. A total of 21 EU/EEA countries reported data on imported dengue, within the period of 2010 – 2015 (inclusive of zero reporting). For our analysis, we considered each imported case, its country of infection (as source country), the reporting country (as destination country) and the reporting month.

### Air travel data

To describe the flow of individuals into Europe, we obtained the passenger air travel data for 2010 – 2015, from the International Air Transport Association (IATA) [32]. IATA Passenger Intelligence Services (PaxIS) data, is the most comprehensive airline passenger’s data available today. Data includes the complete passenger itineraries, true origins and destinations, route segments and connecting points. The data contained over 11,996 airports in 229 different countries and their territorial dependencies, with calibrated passenger travel volumes for each route at a monthly timescale. The passenger volumes were available at the country level, i.e. the total number of passengers traveling from each country worldwide. We used these data to construct a monthly directional passenger flow from all countries worldwide with a final destination in Europe (also accounting for all connecting flights). This passenger flow was inclusive of flow in-between European countries.

### Connectivity indices

We assembled seven indices representing factors potentially mediating the importation risk of dengue from a source country. These indices are decomposed into components representing the ‘source strength’ (the risk of dengue infection); and the transport and importation potential (the connection between the source country and the potential destination country in Europe).

Indices 1 and 2, characterize dengue monthly activity and annual seasonality in the source country. These indices were created using worldwide dengue outbreak notifications data from DengueMap (a unified data collection tool, that brings together disparate dengue reports of local or imported dengue cases from official, newspapers and other media sources globally) [33]. **Index 1** is defined as having one or more confirmed cases in a given month (January-December) in the years of 2010 through 2015, **index 2** is defined as having dengue activity (i.e. notification of one or more confirmed cases) in a given month in two or more years from 2010 through 2015 [34]. **Index 3** is the annual dengue incidence estimates of source country adjusted for the country’s population, obtained from the Institute for Health Metrics and Evaluation (IHME) [35, 36]. **Index 4** is the geographical distance between centroids of source country and destination country, often modelled as a proxy for travel time and predictor for epidemic arrival times [37]. This assumes that proximity to an endemic source increases transport and importation potential to destination. **Index 5** is the epidemic vulnerability of the source country, represented by the recent infectious disease vulnerability index from RAND cooperation [38]. This index identifies countries ‘vulnerabilities to control potential disease outbreaks by assessing a confluence of seven broad country level factors: demographic, health care, public health, disease dynamics, political-domestic, political-international and economic [38]. This index assumes that most vulnerable countries might pose a higher risk of infection and transportation. **Index 6** is the income per capita (GDP) of source countries; poor countries with weak economies are associated with poor health outcomes, lesser abilities to detect, prevent and respond effectively to infectious disease. Hence, we assume greater importation risk from poorer source countries. **Index 7** is the total arriving passengers from source country to destination country (i.e. accounting for both direct and connecting flights) which has often been correlated with disease importation [13], with an implicit assumption that infection risk is equal for all source countries. Hence an increase in passengers, in turn, increases the transport and importation potential.

Source strength for all indices was determined by the endemicity of dengue in the source country, while transport and importation potential were modelled based on each unique factor, as detailed in Table 1 below.

**Table 1.**
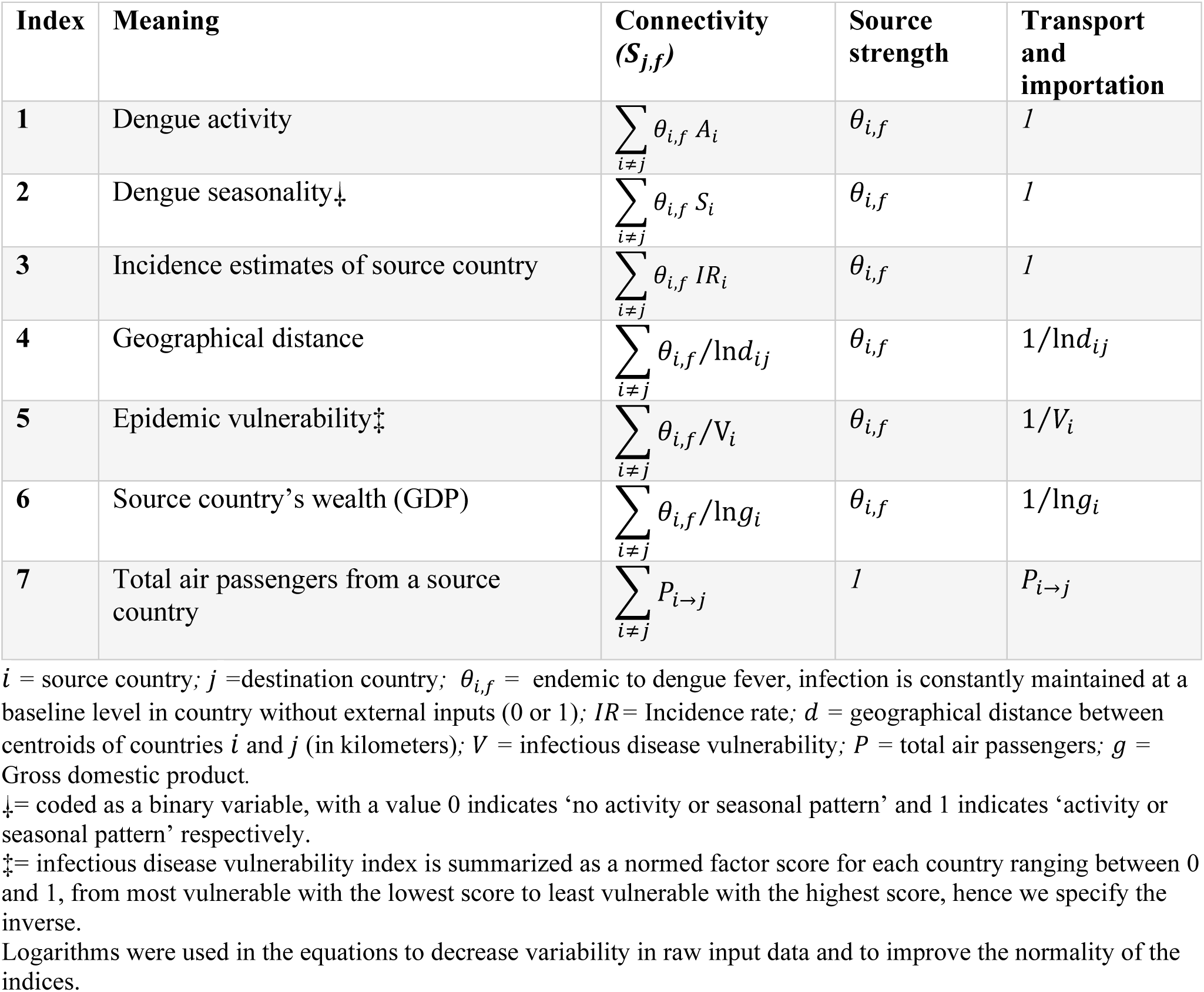
Connectivity indices (S_j,f_)of a focal destination country (j) for the importation of dengue fever (f)

To quantify the variation in dengue importation as explained by our proposed connectivity indices, we fitted a generalized linear mixed effect model (GLMMs) [39] Unit of analysis was the monthly source-destination country combination, with a binary response variable coded to indicate a reported case of imported dengue (1) or not (0). The GLMM was fitted with logit link functions for binomial errors and fixed effects of the connectivity indices and a crossed random effects (intercepts) of the destination country and time step (in a month). We also included a fixed effect for time to control for the following: specific global aviation traffic effects, i.e. increase or decrease in traffic to a specific country, due to a major event (e.g. traffic increase to London, during the 2012 summer Olympics), and to account for under-reporting or no report for countries in specific months. To improve normality, the continuous fixed effects variables were transformed using the log(*x* + 1) function, then centered on zero, and standardized to unit variance, before model fitting. Model fits were evaluated by calculating the marginal and conditional GLMM *R*^2^ [40]. Likewise, model post-estimation using model diagnostic measures and residuals plots were evaluated using the DHARMa residual diagnostics for hierarchical models [41].

### Dependency network

Next, to explore the influence of the air transport network structure on the variation of dengue importation, we incorporate the dependency network approach. This is a system level analysis of the activity and topology of the air transport network to investigate the capacity of a country to influence or be influenced by, other countries within the network by virtue of their connections [42-44]. The specificity of network influence on dengue importation risk is that the connectivity between a source country *(i)* and a destination county *(j)* is not analyzed in isolation, but in consideration for the effects of *z*, where *z*, is the other countries within the network structure. The implication of this network structural view is that the connection between *i* and *j* is also dependent on the connections between *i* and *z* and between *j* and *z*. Therefore, the distributed heterogeneity in the connections is characterized by interdependencies within the network and must be accounted for statistically [25, 45].

Using IATA passengers air travel data, we constructed 72 weighted directed networks, to represent the monthly flow of air passengers into Europe, from 2010 to 2015. For each network, countries are represented by a node while edges represent the flow of passengers between pairs. The network graph is denoted by *G*_*m*_ *= (V, E)*, where *V*_*G*_ is a set containing all the nodes (or vertices), while *E*_*G*_ contains all the edges, with *m* indicating the month. Edges are denoted as *e*_*i,j*_, where *i* is the source and *j* is the destination of a travel route represented by the edge. For each connected pair of nodes *i* and *j*, the edge was weighted with the total number of passengers from *i* to *j* given by *W*_*ij*_ (Fig 1).

**Fig 1.**
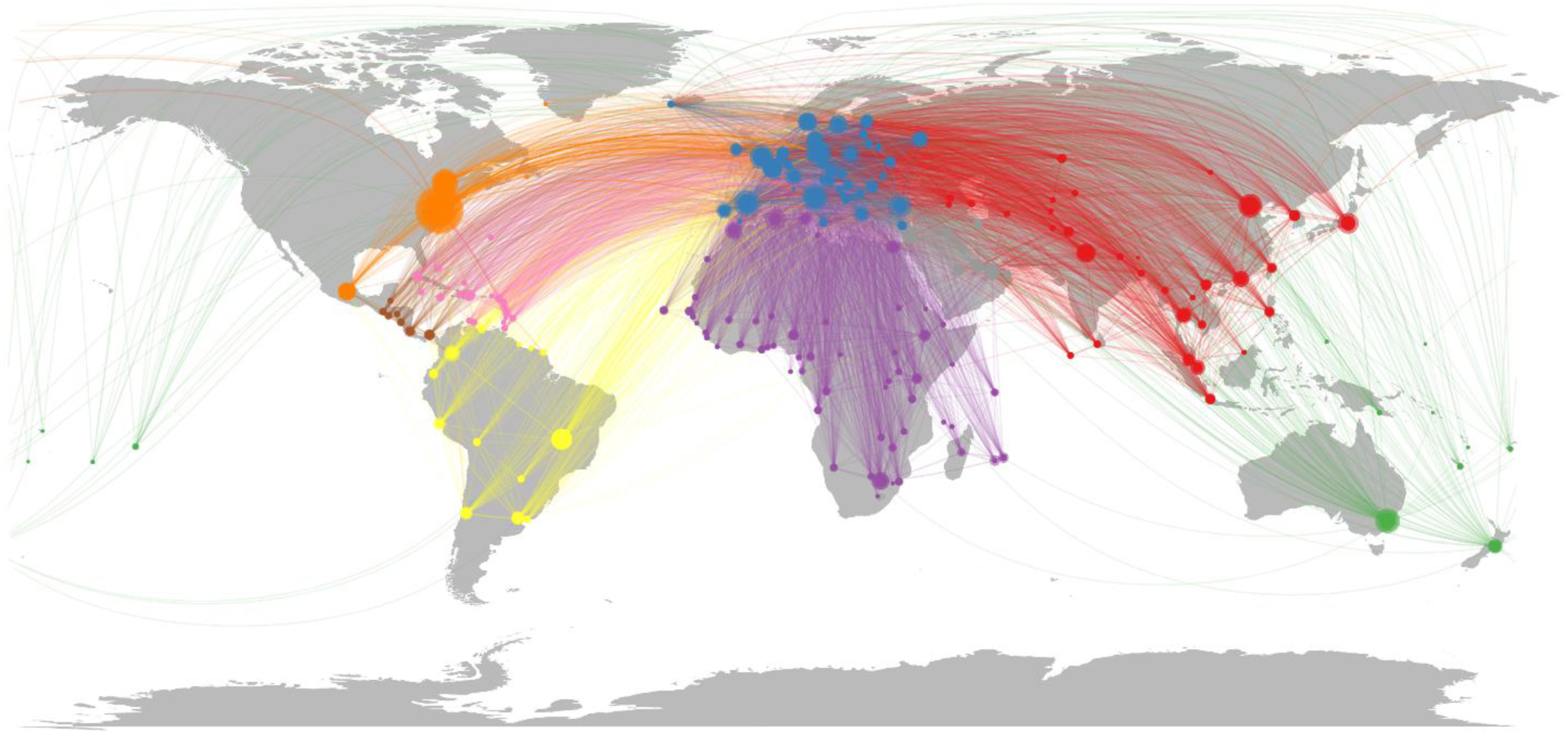
Air transport network. The weighted, directed network, constructed from IATA passenger’s data with a final destination in Europe. Each node is one country, and the size of a node is proportional to the average number of passengers in a month.

The role of a node in the network and its likelihood to influence the transport and importation potential of dengue was then characterized by the following centrality measures (Table 2):

**Table 2.** Centrality measures for a central node (adapted from [45])

|  | Centrality measure | Characteristics of a central node | Equation |
| --- | --- | --- | --- |
| 1 | Degree | Connected directly with many other nodes | $DC_i = s_i = \sum_{j \neq i} W_{ij}$ |
| 2 | Betweenness | Lies on many shortest paths linking other pairs. The probability that communication from $p$ to $g$ will go through $i$ | $BC_i = \sum_{p \neq i, p \neq q, q \neq i} \frac{g_{pq}(i)}{g_{pq}}$ |
| 3 | Closeness | Short communication path to other nodes, a minimal number of steps to reach others | $CC_i = \frac{N}{\sum_j l_{ij}}$ |
| 4 | Eigenvector | Connected (directly and/or indirectly) to many other nodes and/or to other high-degree nodes | $EC_i = \frac{1}{\lambda} \sum_j A_{ij} v_j$ |

$i$ = source country; $j$ =destination country; $s$ = node strength (i.e. the sum of all edge weights attached to a node); $W_{ij}$ = weight of the edge between nodes $i$ and $j$ ; $g_{pq}$ = the number of shortest-paths between nodes $p$ and $q$ ; $g_{pq}(i)$ = the number of shortest-paths between nodes $p$ and $q$ (other random nodes in the network), which pass through $i$ ; $N$ = number of nodes in a network; $l_{ij}$ = distance of the shortest between nodes $i$ and $j$ , here weighted distance is given by the inverse of the number of passengers in the corresponding edge; $A$ = adjacency matrix of the weighted network; $\lambda$ = leading eigenvalue of $A$ ; $v$ = leading eigenvector of $A$ ;

#### Degree centrality

The number of links or connections that a node has, this assigns a score based on the number of nodes within the network, that an individual node is connected to [42, 45]. The higher a node‘s degree, the more it associates with neighboring nodes, potentially increasing its transport potential or increases its vulnerability to importation (in the case of a destination country).

#### Betweenness centrality

Measures the number of times a node lies on the shortest path between other nodes in the network [42]. A node with high betweenness metric is expected to have a higher transport potential, as it bridges between different nodes that are not directly connected. Alternatively, destination countries with high betweenness are at higher risk of importation.

#### Closeness centrality

measures how close (in terms of topological distance) a node is with respect to all other nodes [46]. The topological distance between any pair of connected nodes is given by the inverse of the number of passengers in the corresponding edge. Therefore, the higher the number of passengers in a given edge, the shorter the distance between them, the faster an infection gets transported or imported.

#### Eigenvector centrality

The basic idea of the eigenvector centrality is that a node’s centrality is determined by the combination of its connections and that of its neighbors [47, 48]. A node will have high eigenvector centrality if it has strong connections with other highly connected nodes. In our context, if a node is connected to other highly influential nodes, the higher its transport potential or the higher its vulnerability to importation from a random source in the network.

To quantify the influence of the air transport network topology, we employ two different modelling approaches. Firstly, we account for the network structure in our connectivity framework, by including the centrality measures of the source country. We tested whether GLMMs based on connectivity indices and the air network centrality measures outperformed the base model based on the connectivity indices alone. GLMMs were fitted with the same random effect structure as above. All possible combinations of centrality measures were considered, resultant models were compared based on their fixed effects ΔAIC and marginal GLMM *R*^2^.

Secondly, we model the network topology of the focal destination country as a dependent variable. This assumes that dengue importation into a focal destination country could be directly related to the country’s position in the air transport network. For this model, centrality measures were fitted as fixed effects, with crossed random effects (intercepts) of each node (i.e. each destination country) and time step (m). As above, all possible models were fitted and compared based on their fixed effects ΔAIC and marginal GLMM *R*^2^.

All analysis and plots were performed in R software, version 3.5.2 [49], using the following libraries: glmmtmb [50], igraph [51], tnam [52], DHARMa [41], sjstats [53], ggplot2 [54], circlize [55] and their varying dependencies.

## Results

Among the 7,277 imported dengue cases reported in Europe from 2010 to 2015, 4,112 (57.0%) cases had known travel history, i.e. source country of infection. The cases with the known source were imported into 21 European countries, from 99 different countries distributed across all global regions. 62.1% of reported cases originated from South-East Asia Region, with Thailand, Indonesia, India, and Sri Lanka being the major source of import; 18.0% originated from Region of the Americas, with most cases coming from Brazil; 12.1% from the Western Pacific Region, where Vietnam and the Philippines were the major import source. The data showed that imported cases were most frequently reported in Germany, Sweden, United Kingdom, Italy and Norway (Fig 2).

**Fig 2.**
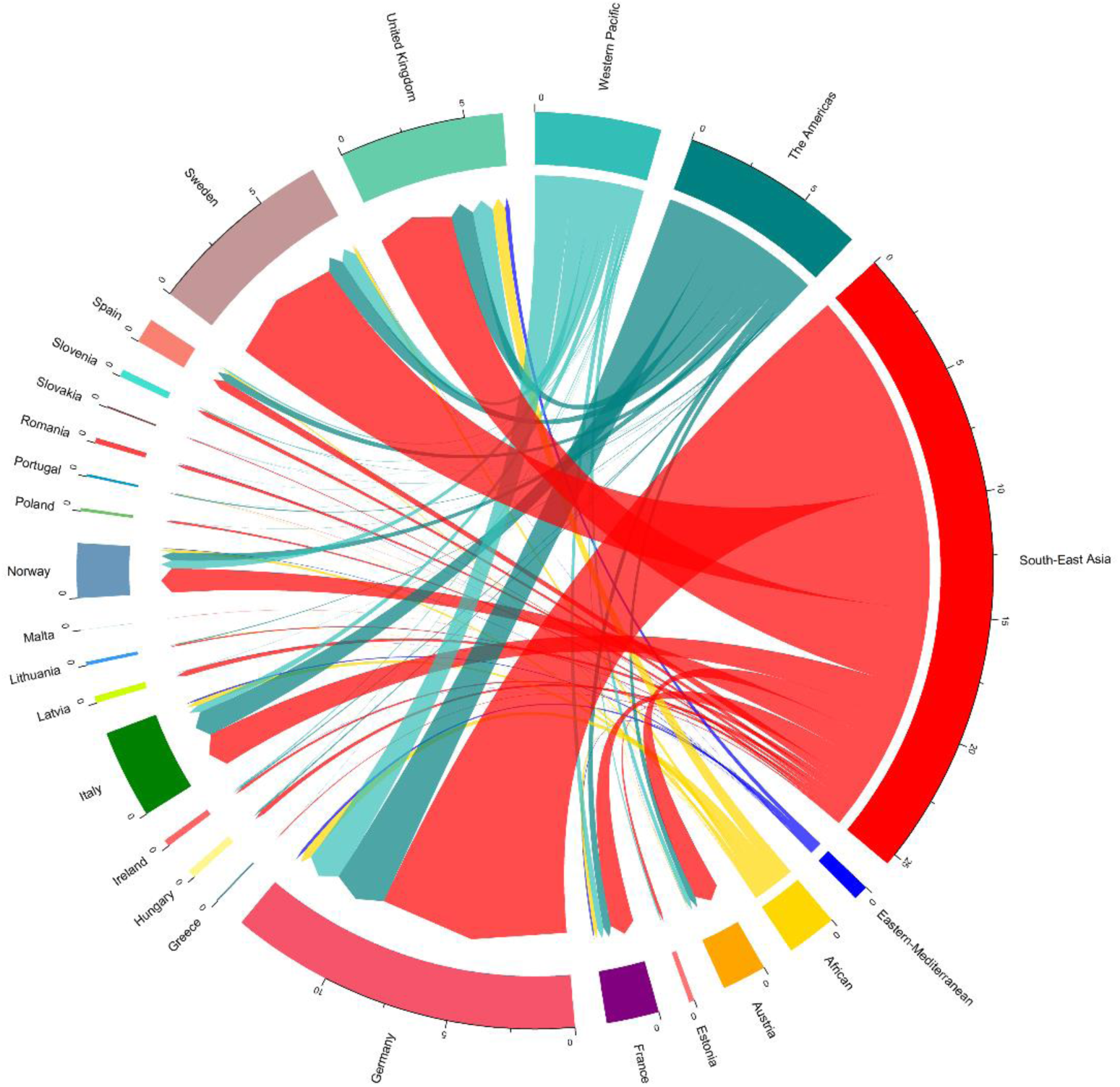
Distribution of imported dengue cases by the destination country and the source region. Destination countries are represented by 21 European countries. Source region represents, 99 different countries distributed across all global. Source countries were grouped into regions for visual representation, with region grouping defined by the WHO Member States region definition. WHO Member States are grouped into 6 WHO regions: African Region, Region of the Americas, South-East Asia Region, European Region (not included in source countries), Eastern Mediterranean Region, and Western Pacific Region [56].

An annual average of 436 million passengers entered Europe from other countries worldwide from 2010 to 2015. Of the total number of passengers arriving from regions outside of Europe, 44% originated from region of the Americas, with higher traffic from the United states, Canada and Brazil; 19% from the Eastern Mediterranean, with higher traffic from

Morocco, United Arab Emirates and Tunisia; 15% from the Western Pacific with higher traffic from Thailand, India and Hong Kong; 12% from the African region, with higher traffic from Algeria, South Africa, Nigeria; 10% from South-East Asia and. A country-level passenger in-flow from WHO Member States regions is shown in Fig 3. High traffic inflow was most common in the United Kingdom, France, Germany, Italy, and Spain. Most of the identified dengue importation hot spots were characterized by a high influx of air passengers, with a tendency for an increase in importation risk. However, the high influx of passengers does not necessarily lead to a high number of imported cases of dengue, as total number of arriving passengers was weakly correlated with the number of imported cases (Spearman’s ƿ = 0.13, p=<0.01, S1 Fig).

**Fig 3.**
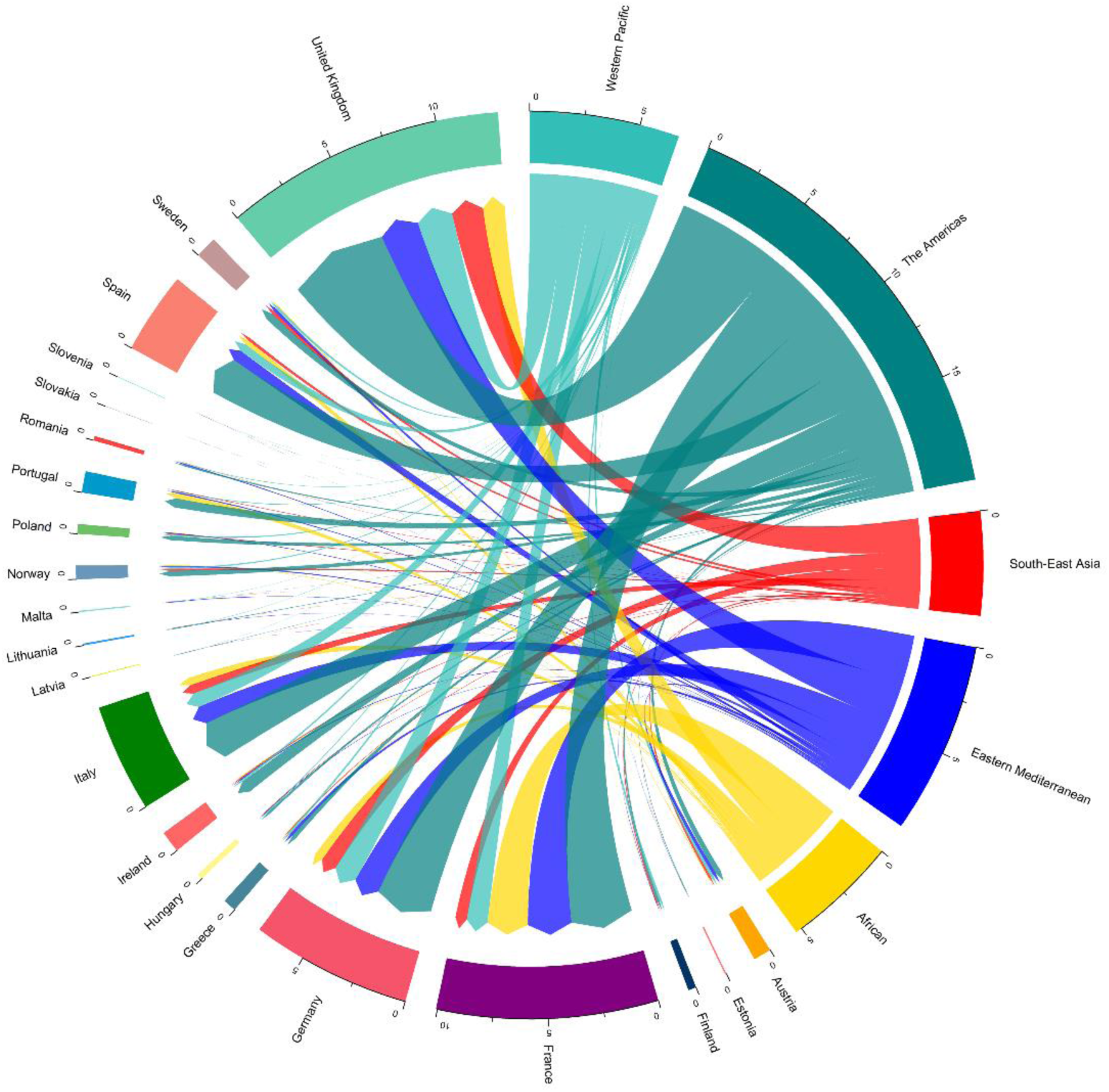
Distribution of air passengers arriving into Europe from WHO regions in 2010 – 2015. Destination countries are represented by 21 European countries. Source region represents, 99 different countries distributed across all global. Source countries were grouped into regions for visual representation, with region grouping defined by the WHO Member States region definition. WHO Member States are grouped into 6 WHO regions: African Region, Region of the Americas, South-East Asia Region, European Region (not included in source countries), Eastern Mediterranean Region, and Western Pacific Region [56].

### Connectivity indices and dengue importation

Our first model investigated the relationship between dengue importation and the connectivity indices. Table 3 presents the results of the GLMM model with estimated coefficients, odds ratios and 95% confidence intervals for each connectivity index. The estimated coefficients represent the relative influence of each connectivity index on the risk of dengue importation between source–destination country combinations. In this model, dengue importation was significantly associated with the following connectivity measures: dengue activity, seasonality, incidence rates, geographical distance, the wealth of source country and arriving passengers. Evaluating the model independently of random effects variance component, these connectivity indices account for 48% (marginal GLMM *R*^2^ of 0.478) of the variation in dengue importation patterns. Overall the GLMM model accounted for 74% (conditional GLMM *R*^2^ of 0.740) of the variation in dengue importation into Europe. The model fit was adequate, the simulated residual diagnostics test for overall uniformity showed no evidence of model misspecification.

**Table 3.**
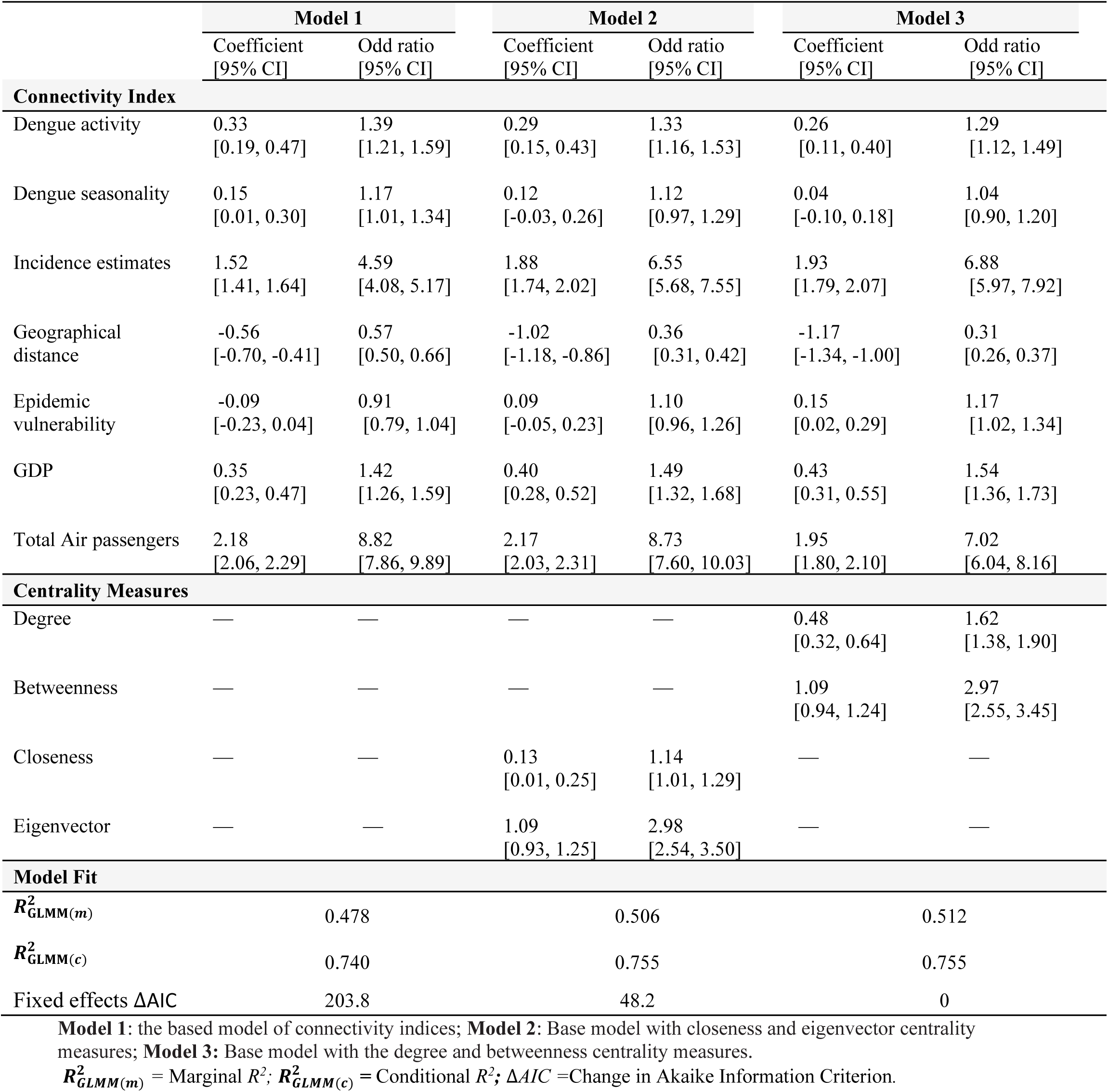
Comparison of generalized linear mixed models predicting dengue importation in Europe, using the connectivity indices and network autocorrelation (i.e. centrality measures).

|  | Model 1 |  | Model 2 |  | Model 3 |  |
| --- | --- | --- | --- | --- | --- | --- |
|  | Coefficient<br>[95% CI] | Odd ratio<br>[95% CI] | Coefficient<br>[95% CI] | Odd ratio<br>[95% CI] | Coefficient<br>[95% CI] | Odd ratio<br>[95% CI] |
| **Connectivity Index** |  |  |  |  |  |  |
| Dengue activity | 0.33<br>[0.19, 0.47] | 1.39<br>[1.21, 1.59] | 0.29<br>[0.15, 0.43] | 1.33<br>[1.16, 1.53] | 0.26<br>[0.11, 0.40] | 1.29<br>[1.12, 1.49] |
| Dengue seasonality | 0.15<br>[0.01, 0.30] | 1.17<br>[1.01, 1.34] | 0.12<br>[-0.03, 0.26] | 1.12<br>[0.97, 1.29] | 0.04<br>[-0.10, 0.18] | 1.04<br>[0.90, 1.20] |
| Incidence estimates | 1.52<br>[1.41, 1.64] | 4.59<br>[4.08, 5.17] | 1.88<br>[1.74, 2.02] | 6.55<br>[5.68, 7.55] | 1.93<br>[1.79, 2.07] | 6.88<br>[5.97, 7.92] |
| Geographical distance | -0.56<br>[-0.70, -0.41] | 0.57<br>[0.50, 0.66] | -1.02<br>[-1.18, -0.86] | 0.36<br>[0.31, 0.42] | -1.17<br>[-1.34, -1.00] | 0.31<br>[0.26, 0.37] |
| Epidemic vulnerability | -0.09<br>[-0.23, 0.04] | 0.91<br>[0.79, 1.04] | 0.09<br>[-0.05, 0.23] | 1.10<br>[0.96, 1.26] | 0.15<br>[0.02, 0.29] | 1.17<br>[1.02, 1.34] |
| GDP | 0.35<br>[0.23, 0.47] | 1.42<br>[1.26, 1.59] | 0.40<br>[0.28, 0.52] | 1.49<br>[1.32, 1.68] | 0.43<br>[0.31, 0.55] | 1.54<br>[1.36, 1.73] |
| Total Air passengers | 2.18<br>[2.06, 2.29] | 8.82<br>[7.86, 9.89] | 2.17<br>[2.03, 2.31] | 8.73<br>[7.60, 10.03] | 1.95<br>[1.80, 2.10] | 7.02<br>[6.04, 8.16] |
| **Centrality Measures** |  |  |  |  |  |  |
| Degree | — | — | — | — | 0.48<br>[0.32, 0.64] | 1.62<br>[1.38, 1.90] |
| Betweenness | — | — | — | — | 1.09<br>[0.94, 1.24] | 2.97<br>[2.55, 3.45] |
| Closeness | — | — | 0.13<br>[0.01, 0.25] | 1.14<br>[1.01, 1.29] | — | — |
| Eigenvector | — | — | 1.09<br>[0.93, 1.25] | 2.98<br>[2.54, 3.50] | — | — |
| **Model Fit** |  |  |  |  |  |  |
| $R^2_{GLMM(m)}$ | 0.478 | | 0.506 | | 0.512 | |
| $R^2_{GLMM(c)}$ | 0.740 | | 0.755 | | 0.755 | |
| Fixed effects $\Delta AIC$ | 203.8 | | 48.2 | | 0 | |

**Model 1:** the based model of connectivity indices; **Model 2:** Base model with closeness and eigenvector centrality measures; **Model 3:** Base model with the degree and betweenness centrality measures.
$R^2_{GLMM(m)}$ = Marginal $R^2$ ; $R^2_{GLMM(c)}$ = Conditional $R^2$ ; $\Delta AIC$ = Change in Akaike Information Criterion.

### Influence of air transport network topology

The resulting air transport network from the passenger’s data was a directed graph structure with 229 nodes and 10261 edges. We took the mean of each centrality measure across the 72 weighted networks to obtain a single value for each node. Based on the mean centrality measures, the following source countries were most central in respect to all metrics: Canada, United States, Australia, United Arab Emirates, China and Brazil (Fig 4). The network metrics displayed notable relationships, with moderate-to-high correlations amongst themselves (S2 Fig). Betweenness and Eigenvector centrality pairs were the most highly correlated (Spearman’s ƿ = 1.00, p <.001). Hence to avoid redundancy in model fitting, we added the correlated pairs in separate models. All the network metrics examined were significant predictors of dengue importation. Compared with the base model of the connectivity indices alone, GLMM fits were substantially improved by including the effects of the network topology (Fig 5). The best-fitting model was the model with degree and betweenness centrality measure pair, alongside the connectivity indices (Table 3). The inclusion of these covariates slightly changed the relative influence of the connectivity indices, with the epidemic vulnerability index becoming statistically significant.

**Fig 4.**
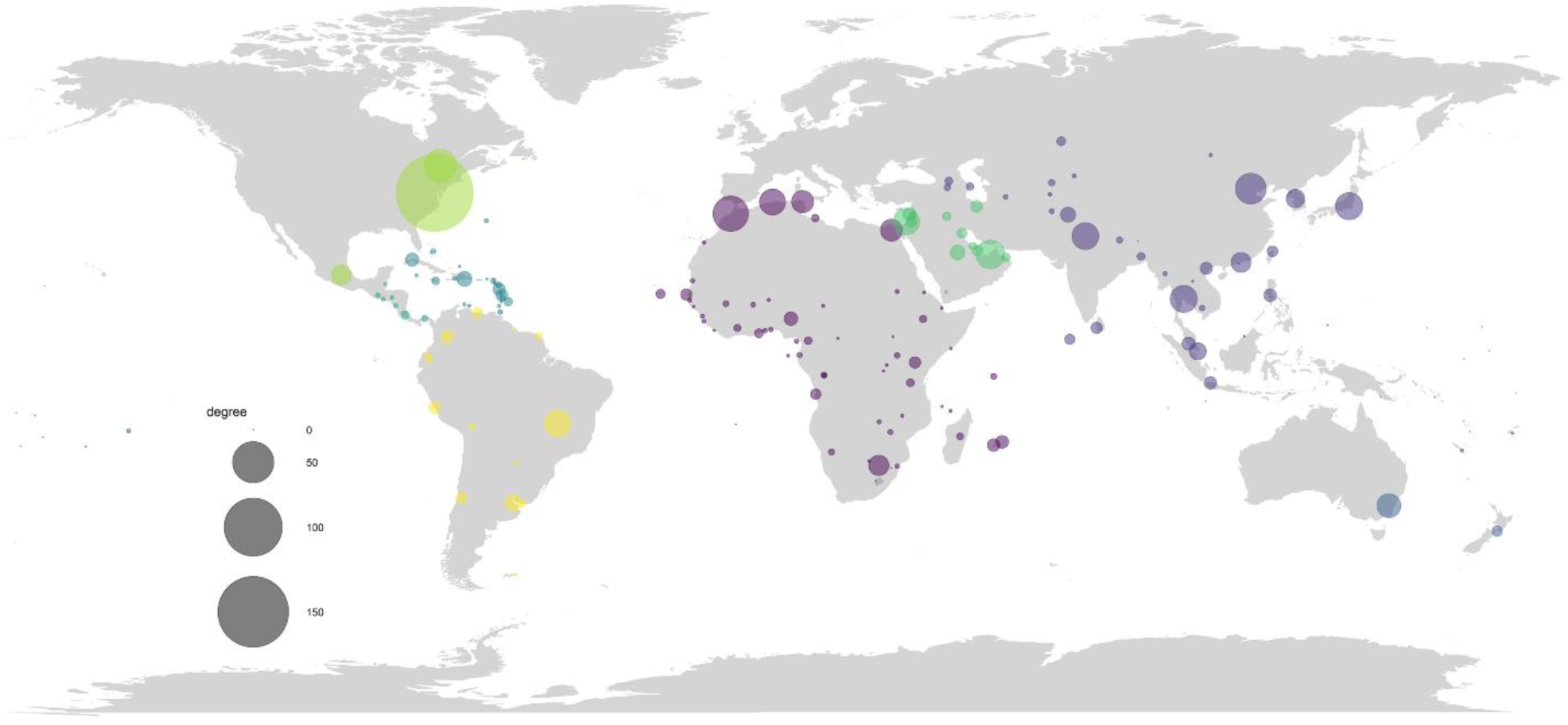
Degree centrality measure of source countries. Each node represents a source country in the air transport network, and the size of a node is proportional to its degree, which is weighted by the average number of passengers with a final destination in Europe. Node color is categorized by region. For visual representation, the actual degree score was scaled down by a factor of 10^4^.

**Fig 5.**
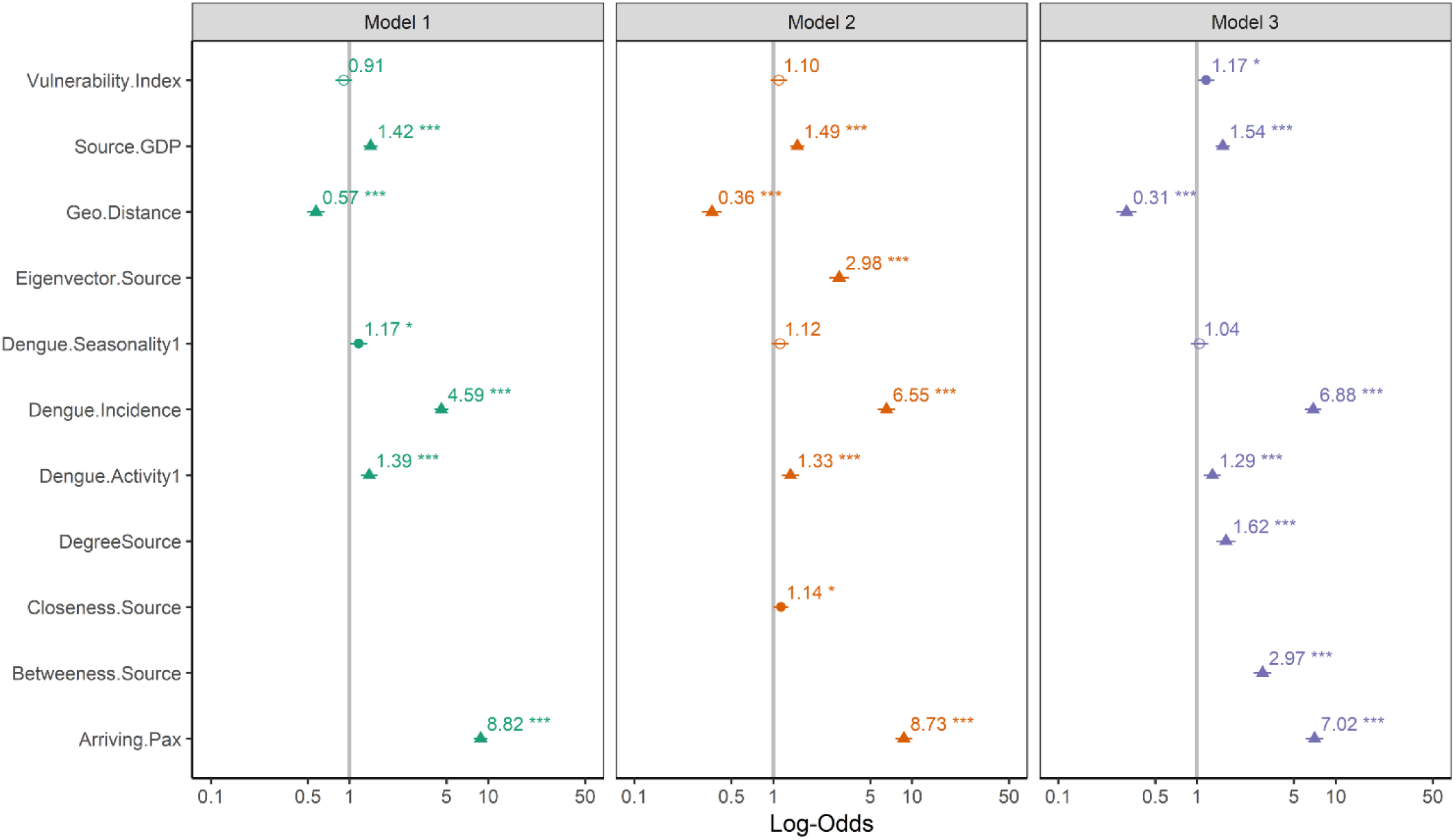
Forest plot comparing the base model and the network autocorrelation models for explaining dengue importation into Europe. Models are as represented in Table 3. Plots of the exponential transformed coefficients estimates, i.e. as an odd ratio. The “no effect” line is set at 1 and denoted by the grey line. Asterisks indicate the significance level of estimates *** = p<0.001; ** = p<0.01; * = p<0.05.

### Network topology as a predictor of importation

Focal destination countries that were most central in the network were the United Kingdom, Germany, Spain, Italy, and France (Fig 6). Therefore, they are most vulnerable to dengue importation from a random source country. The centrality measures were highly correlated amongst themselves, hence it was practically redundant to include all measures in a single model (S2 Fig). We added each centrality measure in separate models, to capture the influence of the unique aspect of each nodal centrality. Each centrality measure was a significant predictor of dengue importation (Table 4). Surprisingly, the eigenvector centrality measure accounted for 70% of the variance in dengue importation (marginal GLMM *R*^2^ of 0.706).

**Table 4.** GLMMs modeling centrality measures of the destination countries as a predictor of dengue importation.

| Network Metrics | Degree | Closeness | Betweenness | Eigenvector |
| --- | --- | --- | --- | --- |
| Coefficient<br>[95% CI] | 1.12<br>[1.06, 1.18] | 1.32<br>[1.22, 1.42] | 1.25<br>[1.18, 1.32] | 2.88<br>[2.65, 3.10] |
| Odd ratio<br>[95% CI] | 3.08<br>[2.90, 3.27] | 3.73<br>[3.37, 4.13] | 3.49<br>[3.26, 3.74] | 17.73<br>[14.12, 22.27] |
| Model Fit |  |  |  |  |
| $R^2_{GLMM(m)}$ | 0.295 | 0.339 | 0.313 | 0.706 |
| $R^2_{GLMM(c)}$ | 0.270 | 0.357 | 0.342 | 0.719 |
| Fixed effect $\Delta AIC$ | 794.7 | 90.2 | 311.5 | 0.0 |

$R^2_{GLMM(m)}$ = Marginal $R^2$ ; $R^2_{GLMM(c)}$ = Conditional $R^2$ ; $\Delta AIC$ = Change in Akaike Information Criterion.

**Fig 6.**
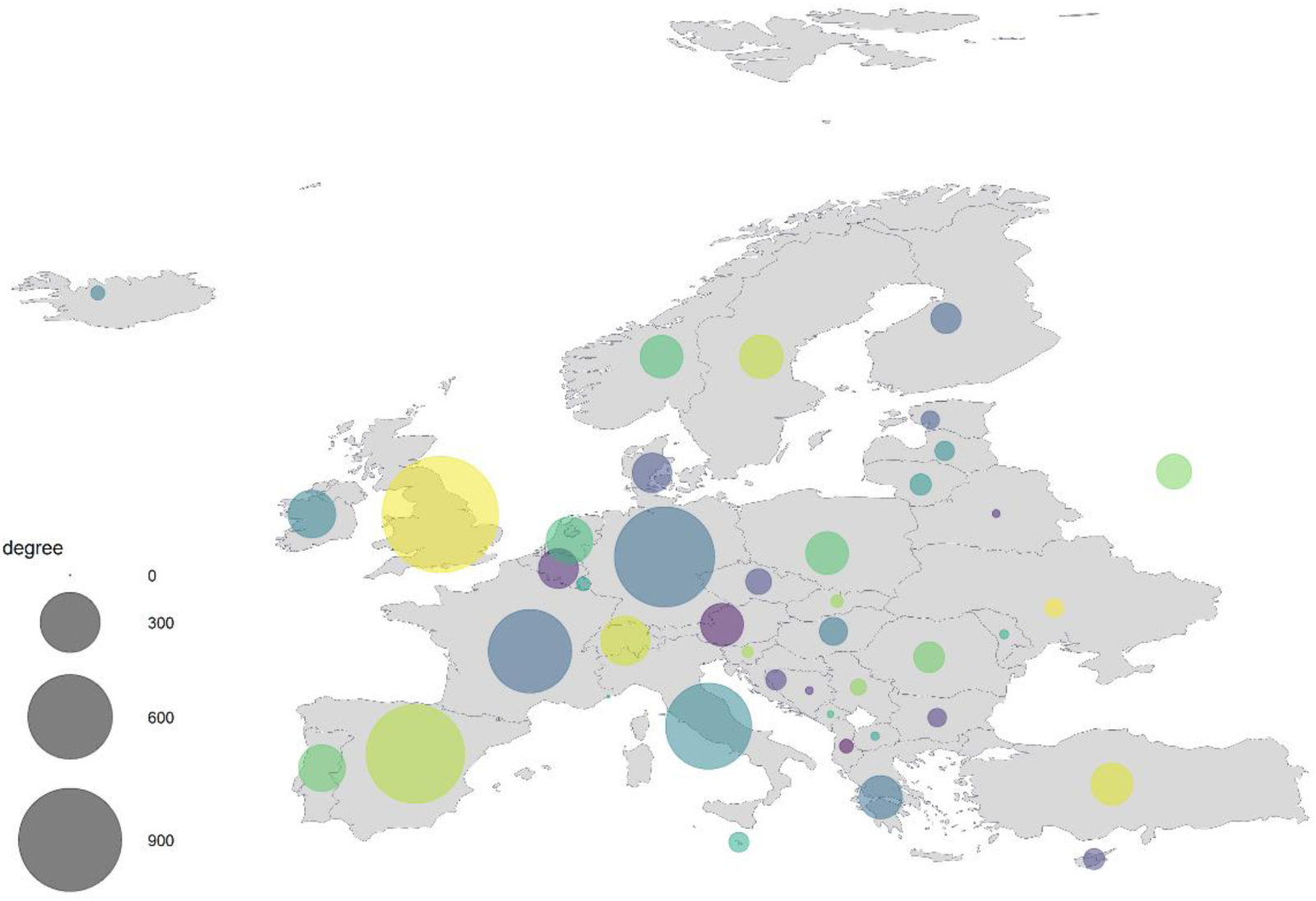
Degree centrality measure of the focal destination countries in Europe. Each node represents a country in Europe, and the size of a node is proportional to its degree, which is weighted by the average number of passengers. For a visual representation, the actual degree score was scaled down by a factor of 10^4^.

## Discussion

The importation of dengue into non-endemic regions is primarily initiated by global human mobility, hence it is critical to understand the dynamics of the transmission pathways based on the mobility networks. Here, we applied a refine network connectivity approach to model the importation of dengue into Europe. Our analysis accounted for factors that mediate the risk of importation from a source country through their effects on source strength and transport potential. In addition, we considered the influence of the air transport network topology on the importation risk from a source and the propensity of a destination country to receive an imported case. Our analysis demonstrated that the co-dynamics of the connectivity indices and the network topology explained more than 70% of the variance in dengue importation patterns. Likewise, the topologically positioning of a focal destination county in the network, played a key role in the importation patterns. These results contribute to our understanding of the transmission pathways of dengue importation and the role of the dynamical processes of the air transport network.

A major focus of our study was the understanding of importation patterns of dengue in Europe, with consideration to source strength and the characterized air transport network. Air passengers’ data and transport connections have been used to infer relationships with imported cases of dengue [13, 57]. However, with the growing complexity of global mobility and transportation links, it is increasingly difficult to model importation solely on the crude aggregate statistics of air traffic [16, 58]. Air traffic connections between source and destination account for a fraction of required modeling parameters for general patterns of importation, other epidemiological and anthropogenic parameters need to be accounted for. A fundamental understanding of other mediating factors and transportation dynamics is required to achieve a more reliable predictive model.

Our analysis addresses this by endowing each source with a specific pattern of connectivity that mediates, its strength (risk of infection), transport and importation potential. Source strength was the risk of infection derived from the endemicity of dengue in a source country, distinguishing endemic and non-endemic countries. Heterogeneities in source strength were further characterized by dengue activity and seasonality patterns in the source country. As expected, an ongoing activity and seasonal pattern of dengue in a source country significantly increase the transport and importation potential. Transport and importation potential were also modelled uniquely for each source by the confluence of the various connectivity indices, accounting for other mediating factors. This modelling approach is corroborated by other similar studies [13, 23, 59]. With the results matching our a priori expectations, i.e. higher transport potential for source countries with high passenger traffic, high incidence rates, lower economic status, and geographical proximity to a destination country.

A new feature of interest in our analysis was the utilization of the infectious disease vulnerability index as an epidemiological factor. The vulnerability index presents a robust tool that identifies a country’s ability to limit the spread of outbreak-prone diseases [38]. The combined multifarious nature of this index offers an intuitive understanding of the indigenous vulnerability of a source country. For our analysis, we modelled source strength, transport, and importation potential to increase with higher vulnerability, as expected most vulnerable countries poses a greater risk (model 3). Although our study was focused on the earliest stages of a network mediated epidemic, the inclusion of this index for focal destination country could provide insights for modeling the establishment potential of an imported case of dengue. With the assumption, that recipient countries with higher vulnerability might present greater dissemination and establishment risk from an imported case, assuming appropriate vector presence [60].

Our analysis went further to incorporate the dependency network approach to account for the influence of the air transport network topology on dengue importation. Utilizing centrality measures to quantify the influence of connection topology and the dynamical processes of the network to influence the importation of dengue [24]. We applied two different network analysis modelling approach, to quantify the unique contributions of each topological descriptor to dengue importation. The first approach incorporates network autocorrelation from the source country’s centrality measures, within the GLMM framework of the connectivity indices. The addition of the centrality measures within the modeling framework addresses the issue of covariance driven by the network structure. All the network descriptors were significant predictors of dengue importation, however the combined effect of degree and betweenness centrality were the most influential. This result suggests that source countries that are highly connected (having multiple air routes into Europe) and act as connecting links to others countries (large airport hubs connecting other countries), intuitively have higher transport and importation potential, as they have the capacities to quickly connect with the wider network.

The second approach investigates if there is evidence of a correlation between the centrality measures of a destination country and its propensity to receive an imported case. We applied this approach as a valuable measure of a direct relationship between the air transport network structure and dengue importation into Europe [25]. Similar to the above results, all the network metrics were strong predictors of the variation in dengue importation. However, the eigenvector centrality, was the most fitting single predictor, explaining over 70% variance in dengue importation. These results suggest that the risk of dengue importation for a country (in Europe) can be largely explained by its position in the air transport network. Meaning countries have higher tendency for an imported case as a result of having more direct ‘one hop’ connections with high passengers traffic (as measured by degree centrality); having large airport hubs, bridging other countries (betweenness centrality); being effectively closer to other countries because of large passenger traffic (closeness centrality); and having multiple direct and indirect connections to other higher connected countries (eigenvector centrality). Overall, these results are particularly valuable in identifying countries in Europe that needs to prioritize investment in real-time surveillance systems, as a health security measure [35, 61], due to their increase propensity to receive an imported case.

In summary, our paper presents a refined approach to the modelling of a network-mediated epidemic for dengue fever. The connectivity indices presented here captures the variation in source strength, transport, and importation potential, by accounting for other mediating socio-economic and anthropogenic factors. These indices are an amenable representation of real-world risk factors but offer a different approach for analyzing the connectivity dynamics of network-mediated importation of dengue. Our analysis went further to characterize the role of the air transport network topology in the dynamics of dengue importation into Europe. By investigating the network autocorrelations influencing transport potential from source countries and the network positioning of the destination country as a predictor of importation. Our analyses show that the connectivity indices and dynamical processes of the air transport network are strong predictors of dengue importation in Europe. Therefore, the network connectivity modelling approach could be useful in predicting source countries, importation patterns and destination countries with greater risk of dengue. Thereby, allowing for preemptive strategies to mitigate the impacts of imported cases in a timely, accurate and cost-effective manner [62, 63]. Finally, this modeling approach could serve as a pivotal prerequisite for the development of an early warning surveillance system to monitor and forecast the spread of dengue fever.

## Data Availability

Disease (dengue) data are available by requested from European Centre for Disease Prevention and Control (ECDC); The air travel data used in this study are owned by the International Air Travel Association(IATA), limited access to this data was furnished under a non-disclosure agreement by International Air Travel Association (IATA)- Passenger Intelligence Services (PaxIS), strictly for the purpose of a doctoral thesis. Sources of all other relevant data are referenced within the paper.

https://www.iata.org/services/statistics/intelligence/paxis/Pages/index.aspx

https://www.ecdc.europa.eu/en/publications-data/european-surveillance-system-tessy

